# Machine Learning for Urinary Tract Infection Prediction in Emergency Departments: An Explainable Approach

**DOI:** 10.64898/2025.12.13.25342059

**Authors:** Ata Dönmez

## Abstract

Urinary tract infections (UTIs) represent a substantial burden in emergency department (ED) settings, where diagnostic delays and the limitations of traditional clinical assessments often result in suboptimal treatment decisions. This study develops an interpretable machine learning framework to enhance real-time UTI prediction accuracy.

We analyzed a retrospective dataset of 80,387 ED patient encounters from four institutions (2013–2016), encompassing 220 clinical variables. Four machine learning algorithms, Decision Tree, Random Forest, Logistic Regression, and XGBoost, were trained and evaluated. Model interpretability was achieved through SHapley Additive exPlanations (SHAP) analysis. Performance was assessed using area under the receiver operating characteristic curve (AUC), sensitivity, and specificity, both overall and across multiple patient subgroups.

XGBoost demonstrated the highest overall performance with an AUC of 0.90 for general UTI prediction and 0.97 for complicated UTI identification. The reduced 20-feature XGBoost model achieved similar performance to the full feature model. SHAP analysis indicated that urinalysis markers, including leukocyte esterase, nitrites, and bacterial count, were the primary predictive features, with additional contributions from age, vital signs, and selected comorbidities. The model maintained robust performance across demographic and clinical subgroups, with AUC values ranging from 0.88 to 0.92.

This study presents a clinically viable, explainable machine learning framework that addresses critical gaps in ED-based UTI diagnosis by combining high predictive accuracy with transparent feature attribution, reduced-feature implementation, and detailed subgroup and benchmark analyses.

## 1. Introduction

Urinary tract infections constitute one of the most prevalent infectious conditions presenting to emergency departments, affecting millions of patients annually and generating substantial healthcare expenditures estimated in the billions of dollars[1]. The diagnostic challenge in acute care settings arises from a fundamental temporal disconnect. While urine culture remains the gold standard for UTI confirmation, results typically require 24 to 48 hours to process[2]. Consequently, emergency physicians must render treatment decisions based on clinical presentation, physical examination findings, and rapid diagnostic tests, an approach associated with considerable diagnostic uncertainty.

This diagnostic ambiguity manifests in two clinically significant error patterns: overtreatment through unnecessary antibiotic administration in culture-negative patients, and undertreatment through missed diagnoses in culture-positive cases presenting with atypical features[3, 4]. Both scenarios carry substantial consequences, including contributions to antimicrobial resistance, adverse drug reactions, inadequate infection control, and increased healthcare utilization.

Recent investigations have demonstrated machine learning’s potential to enhance UTI diagnostic accuracy through systematic analysis of electronic health record (EHR) data. Taylor et al. (2018) established proof of concept by applying multiple machine learning algorithms to a large ED dataset, achieving area under the curve (AUC) values exceeding 0.90 for UTI prediction[5]. However, their work, while methodologically rigorous, primarily evaluated models that functioned as black boxes, generating accurate predictions without providing clinically interpretable explanations for individual classification decisions or detailed subgroup performance.

Limited interpretability represents a critical barrier to clinical adoption. Healthcare providers require not only predictive accuracy, but also transparent reasoning that aligns with clinical knowledge, supports decision accountability, and facilitates appropriate skepticism when model outputs appear incongruent with clinical judgment. The absence of interpretability in many machine learning applications has contributed to reluctance among physicians to integrate such tools into clinical workflows.

Using the same publicly available dataset originally analyzed by Taylor et al.[5], this investigation extends prior work by developing and evaluating an interpretable machine learning framework for ED-based UTI prediction. We explicitly focus on explainability, parsimony, subgroup robustness, and benchmarking against proxy measures of clinician decision-making, to move beyond demonstration of raw predictive performance.

Our specific objectives are threefold.

First, performance evaluation: compare multiple machine learning architectures (Decision Tree, Random Forest, Logistic Regression, XGBoost) for UTI prediction accuracy using comprehensive clinical data, including development of a reduced-feature XGBoost model suitable for streamlined clinical implementation.

Second, interpretability enhancement: implement SHapley Additive exPlanations (SHAP)[6] methodology to provide clinically transparent feature attribution for model predictions and to characterize both global and case-level model behaviour.

Third, subgroup and benchmark analysis: assess model performance consistency across clinically relevant patient subgroups and compare model outputs with proxy measures of clinical decision-making to evaluate generalizability, potential algorithmic bias, and relative performance against current practice.

By integrating predictions with transparent decision explanation and practical implementation considerations, we aim to produce a clinically actionable tool suitable for real-world ED deployment and to provide a reproducible extension of existing work on this dataset.

## 2. Methods

### 2.1 Data Source and Study Population

We utilized a publicly available dataset derived from four academic emergency departments, capturing patient encounters between March 2013 and May 2016[7]. The dataset comprises 80,387 unique ED visits, each characterized by 220 distinct clinical variables. Inclusion criteria encompassed all adult patients presenting to participating EDs who underwent urinalysis testing. The dataset was designed for machine learning applications and contained no missing values across included variables.

### 2.2 Feature Categories and Clinical Variables

Clinical features were organized into seven major categories reflecting comprehensive patient assessment.

| Categories | Variables |
| --- | --- |
| Demographic & Administrative | Age, Gender, Race, Ethnicity, Language Preference, Marital Status, Employment Status, Insurance Classification, Arrival Mode |
| Vital Signs | Temperature, Heart Rate, Systolic Blood Pressure, Diastolic Blood Pressure, Respiratory Rate, Oxygen Saturation, Supplemental Oxygen Requirement, Glasgow Coma Scale |
| Urinalysis | Bacteria, Bilirubin, Blood, Clarity, Color, Epithelial Cells, Glucose, Ketones, Leukocyte Esterase, Nitrites, pH, Protein, Red Blood Cells, Specific Gravity, Urobilinogen, White Blood Cells |
| Physical Examination | Costovertebral Angle Tenderness, Abdominal Tenderness, Abdominal Distention, Guarding, Masses, Rebound Tenderness, Rigidity, Dysuria, Hematuria, Polyuria, Decreased Urinary Volume, Difficulty Urinating |
| Laboratory Values | CBC Parameters, CMP Parameters, Differential WBC Count |
| Medical History | Renal Disease, Diabetes, Urinary Tract Abnormalities, Immunosuppression, Cardiac Conditions, Urological Conditions |
| Current Medications | Antibiotics, Immunosuppressants, Urinary Function Medications, Other ATC Classes |

### 2.3 Data Preprocessing

Given the absence of missing values in the dataset, preprocessing focused on ensuring data integrity and preventing information leakage. We excluded three variables from model training: chief complaint (a free-text field with potential for data leakage), patient ID, and visit ID (non-informative identifiers). Continuous variables were examined for distributional characteristics. Median imputation procedures were prepared but ultimately unnecessary. Categorical variables representing medical conditions and medications were pre-coded using standardized classification systems (CCS and ATC, respectively), and therefore required no further transformation.

### 2.4 Outcome Definition

The primary outcome was laboratory-confirmed UTI, defined as positive urine culture growth of at least 10,000 colony-forming units per milliliter of a single uropathogen or at least 100,000 CFU/mL of mixed organisms. This microbiological standard served as the reference classification for all model training and evaluation procedures.

### 2.5 Machine Learning Algorithms

We implemented four distinct machine learning architectures from sci-kit learn[8] to represent a spectrum from interpretable linear models to complex ensemble methods.

Logistic Regression was included as a linear classifier establishing baseline performance and representing traditional statistical approaches. The regularization parameter (C) was optimized through cross-validation.

The Decision Tree model provided inherent interpretability through hierarchical rule structures. Maximum depth and minimum samples for splitting were tuned to balance model complexity and generalization.

Random Forest, an ensemble method combining multiple decision trees through bootstrap aggregation, was used to capture nonlinear relationships and complex interactions[9]. We optimized the number of estimators and maximum tree depth.

XGBoost (eXtreme Gradient Boosting) is an advanced gradient boosting framework employing sequential tree construction with regularization[10]. This algorithm was selected as our primary model based on its demonstrated success in healthcare prediction tasks and its compatibility with SHAP interpretation methods.

### 2.6 Model Development Strategy

#### Data partitioning

The dataset was randomly divided into training (80%, n = 64,310) and testing (20%, n = 16,077) subsets using stratified sampling to preserve outcome prevalence. The training set was used exclusively for model development and hyperparameter optimization, while the test set remained isolated until final performance evaluation.

#### Hyperparameter optimization

For each algorithm, we conducted systematic hyperparameter tuning using five-fold cross-validation on the training set. Optimal parameters were identified as follows.

Decision Tree: maximum depth 10, minimum samples per split 2.

Random Forest: 200 estimators, unrestricted maximum depth.

Logistic Regression: C 10.0, maximum iterations 1,000.

XGBoost: 300 estimators, maximum depth 3, learning rate 0.2.

#### Feature selection and reduced model

Full models utilized all 217 eligible features after exclusions. In addition, we developed a reduced XGBoost model incorporating the 20 highest-importance variables identified through permutation importance analysis: ua_leuk_negative, ua_nitrite_positive, ua_wbc_small, ua_color_yellow, ua_wbc_large, neutrophils_not_reported, BUN_not_reported, ua_bacteria_many, ua_leuk_small, ua_leuk_large, abxUTI_no, ua_epi_large, ua_bili_negative, ua_clarity_not_reported, ua_bili_large, gender_Female, ua_clarity_clear, absolute_lymphocyte_count_not_reported, ua_urobili_not_reported, and ua_bacteria_marked. This reduced model was designed to approximate full-model performance with a more parsimonious feature set.

### 2.7 Model interpretability framework

To address the need for clinical transparency, we implemented SHAP, a unified approach to interpreting model predictions based on cooperative game theory[8]. SHAP values quantify each feature’s contribution to individual predictions by estimating the change in expected model output when that feature is observed versus absent, averaged over all possible feature combinations.

For the XGBoost model, we utilized TreeSHAP, an optimized algorithm that provides exact SHAP value computation for tree-based models with polynomial rather than exponential computational complexity. SHAP summary plots were generated to visualize global feature importance and the directional impact of feature values on predictions. For case-level interpretation, we examined individual force plots to illustrate how specific feature values contributed to a patient’s predicted probability of UTI.

### 2.8 Performance Evaluation

#### Primary Metrics

Model discrimination was quantified using the area under the receiver operating characteristic curve (AUC-ROC), which measures the probability that the model ranks a randomly selected positive case higher than a randomly selected negative case. We also computed sensitivity (true positive rate) and specificity (true negative rate) at the optimal threshold determined by Youden’s index. Where appropriate, we report AUC values for both overall UTI prediction and complicated UTI detection.

#### Subgroup Analysis

To evaluate model consistency and potential algorithmic bias, we conducted stratified performance evaluation across clinically relevant subgroups:

- Gender (male, female).
- Disposition (admitted to hospital, discharged from the ED).
- UTI complexity (complicated versus uncomplicated cases, with complicated UTIs defined by presence of systemic infection signs, urological abnormalities, or immunocompromise).

#### Clinical Benchmark Comparison

We compared model performance against proxy measures of clinical decision-making derived from the dataset. These benchmarks included documented UTI diagnoses in discharge records and administration of UTI-directed antibiotic therapy, excluding cases with clear alternative infection sources. This analysis enabled comparison between algorithmic predictions and existing clinician behaviour on the same cohort.

### 2.9 Software and Statistical Analysis

All analyses were conducted using Python 3.8 with the following libraries: pandas for data manipulation, scikit-learn for model implementation and evaluation, XGBoost for gradient boosting, SHAP for model interpretation, and matplotlib and seaborn for visualization. Statistical significance was set at α = 0.05, although our primary focus was effect size, particularly AUC differences, rather than p values given the large sample size.

## 3. Results

### 3.1 Cohort Characteristics

| Category / Variable | Value |
| --- | --- |
| Cohort size – Total ED encounters | 80,387 |
| Age – Median age | 53 years |
| Age – Interquartile range | Not reported |
| Sex – Female patients | Approximately 68% |
| UTI status – Culture-confirmed UTI | 18,284 encounters (23%) |
| UTI status – Documented UTI diagnosis (culture-positive) | 36.7% |
| UTI status – UTI-targeted antibiotics (culture-positive) | 79.4% |
| UTI status – UTI diagnosis (culture-negative) | 6.7% |
| UTI status – UTI-targeted antibiotics (culture-negative) | 50.6% |
| Race/Ethnicity – White/Caucasian | 54%–56% |
| Race/Ethnicity – Black/African American | 20%–21% |
| Race/Ethnicity – Hispanic/Latino | 2%–3% |
| Insurance – Commercial | 32%–36% |
| Insurance – Medicaid | 27%–30% |
| Insurance – Medicare | 26%–35% |
| Arrival mode – Private vehicle | 50%–51% |
| Arrival mode – EMS | 31%–37% |
| Arrival mode – Walk-in | 10%–15% |
| Disposition – Hospital admission | 45%–49% |
| Disposition – Discharged | 50%–54% |

### 3.2 Comparative Model Performance

Figure 1 presents receiver operating characteristic curves for all four algorithms. On the held-out test set, XGBoost achieved the highest overall AUC at 0.90 (95% confidence intervals were not available from the original analysis). Random Forest and Logistic Regression both yielded AUC values of 0.89, while the Decision Tree model achieved an AUC of 0.86.

**Figure 1.**
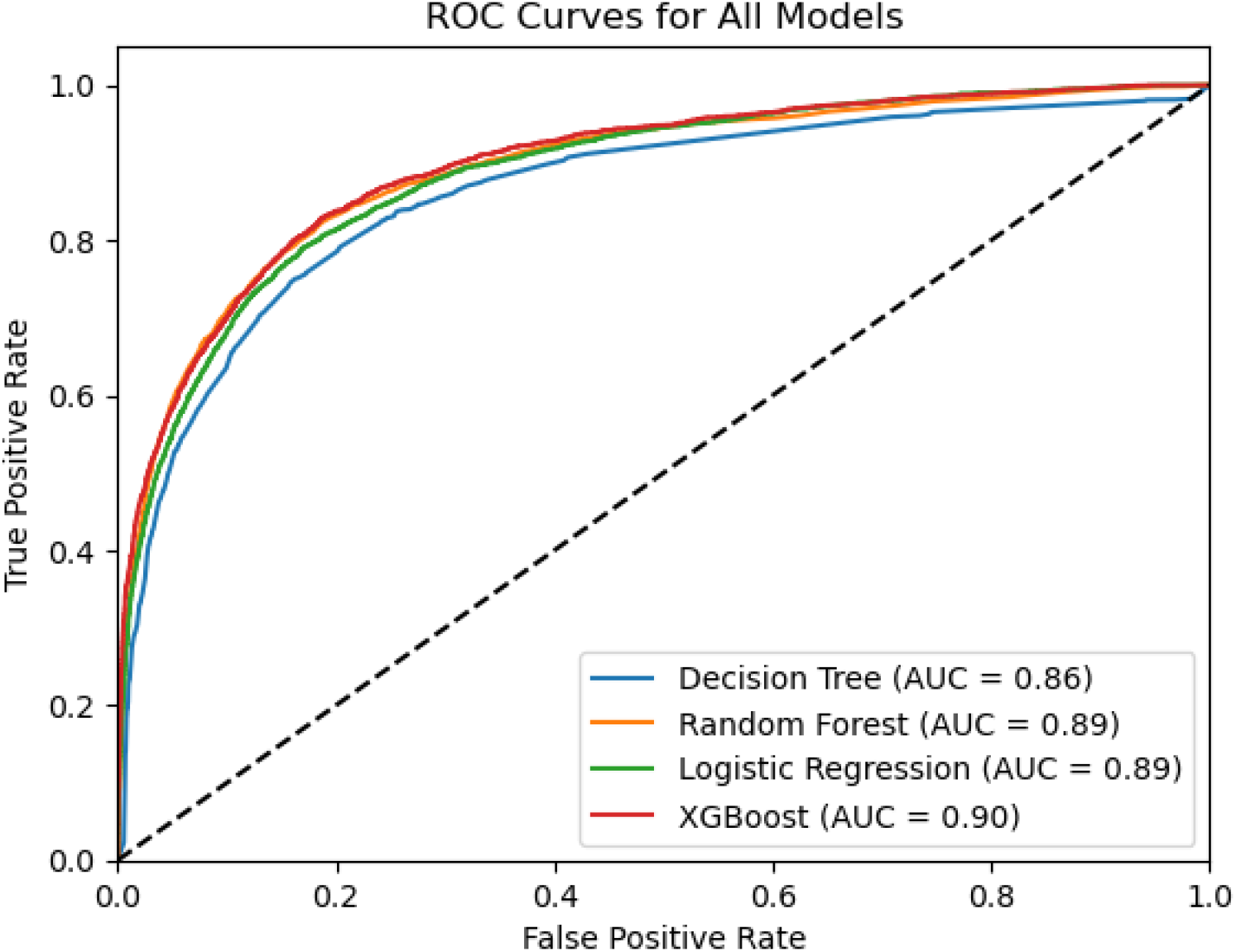
ROC curves comparing the performance of all four models (Decision Tree, Random Forest, Logistic Regression, and XGBoost) for predicting UTI. The diagonal dashed line represents random chance (AUC 0.5).

The reduced XGBoost model, which used only 20 selected features, maintained performance comparable to the full model, with an AUC that differed by less than 0.01 from the 217-feature implementation. Sensitivity and specificity at the Youden-optimal threshold for the full XGBoost model were high; corresponding values for the reduced model were similar.

### 3.3 Feature Importance and Clinical Interpretation

Figure 2 displays the SHAP summary plot for the XGBoost model, showing both global feature importance and directional effects on predicted UTI probability.

**Figure 2.**
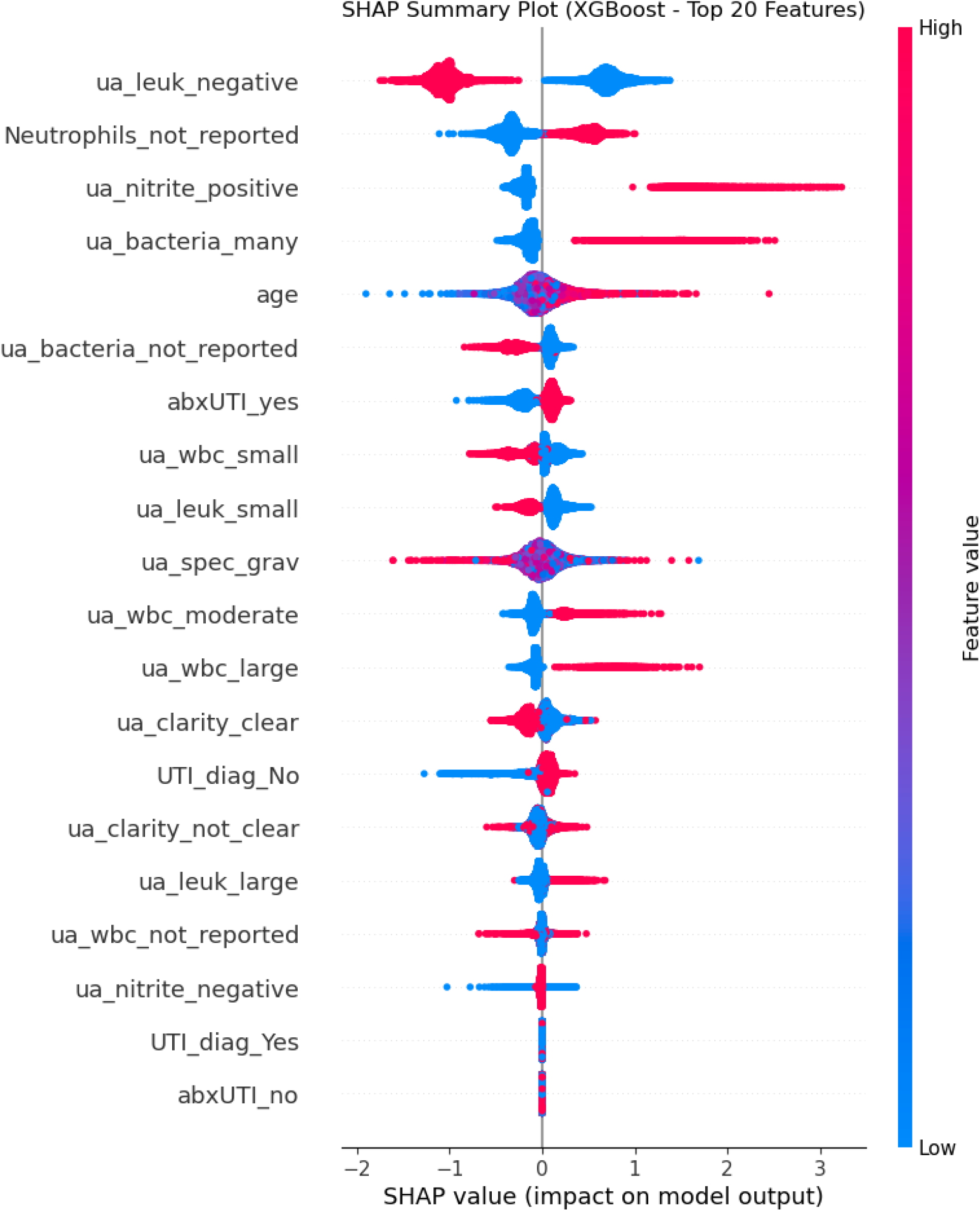
SHAP summary plot for the XGBoost model, displaying the top features

The most influential predictive features were urinalysis components. The presence of nitrites strongly increased predicted UTI probability. Moderate to marked bacteriuria on microscopic examination similarly increased predicted probability. Leukocyte esterase was influential in both directions, with negative results associated with lower predicted probability and positive results (small, moderate, or large) associated with higher predicted probability. Elevated urinary white blood cell counts were associated with higher predictions.

Beyond urinalysis markers, age, temperature measurements, sex, and selected comorbidity and medication variables contributed to model predictions. For example, female sex and higher age values were associated with higher predicted probability. Variables coded as “not reported”, such as neutrophils_not_reported or BUN_not_reported, also appeared among influential features, indicating that patterns in test ordering contributed information to the model.

### 3.4 Performance for Complicated UTI Prediction

Figure 3 illustrates model performance for predicting complicated UTIs, defined as infections with systemic manifestations, structural urinary tract abnormalities, or occurrence in immunocompromised hosts. The XGBoost model achieved an AUC of 0.97 for this subgroup. Corresponding sensitivity and specificity values at the Youden-optimal threshold were higher than those observed for general UTI prediction. The reduced-feature XGBoost model achieved AUC values for complicated UTI that were similar to the full model.

**Figure 3.**
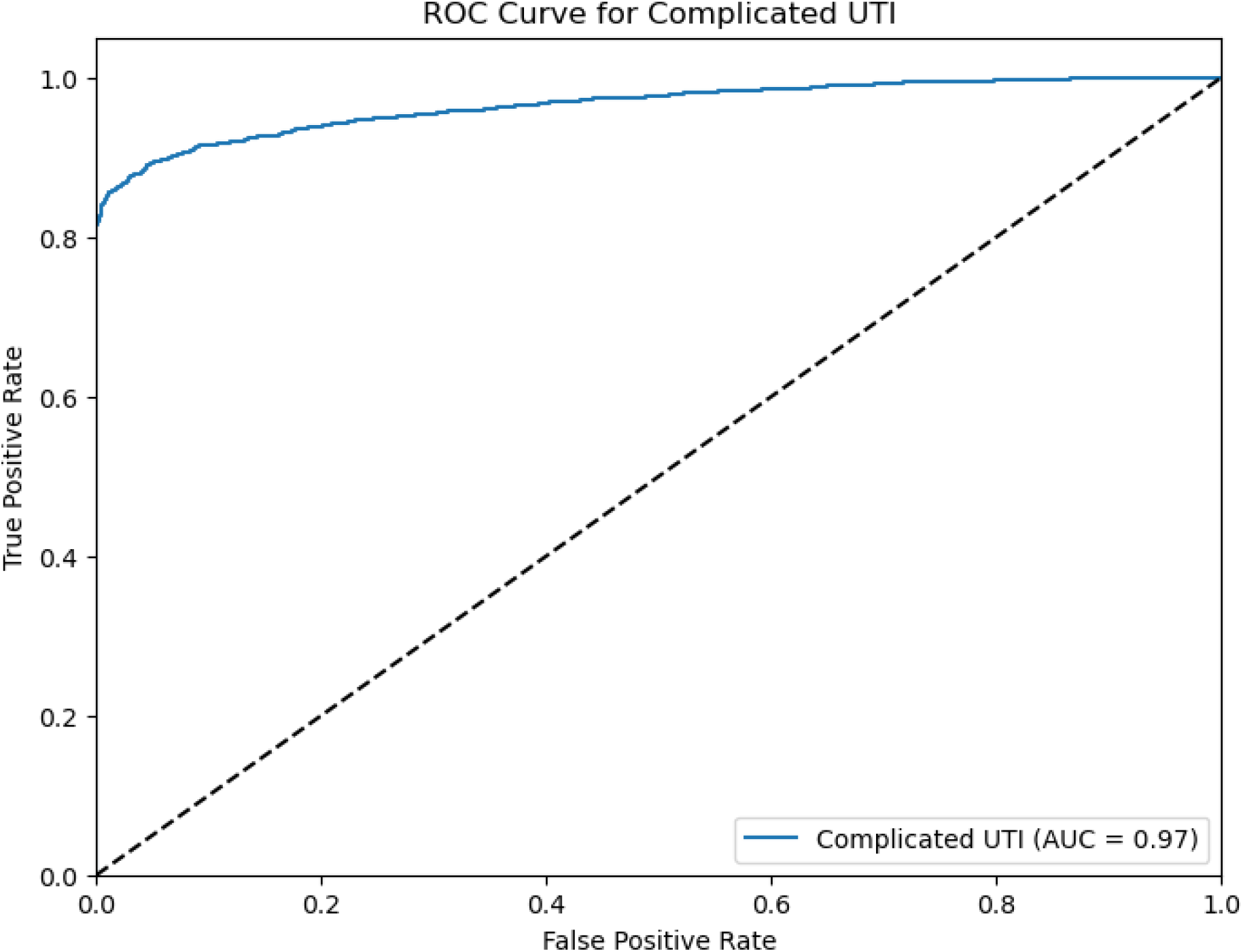
ROC curve for predicting complicated UTIs using the XGBoost model.

### 3.5 Subgroup Performance Analysis

Figure 4 presents stratified ROC curves examining model performance across patient subgroups defined by disposition and gender. The XGBoost model maintained robust discrimination across all groups. For admitted patients, the AUC was 0.91; for discharged patients, AUC was 0.88. For male patients, AUC was 0.92, and for female patients, AUC was 0.88. Sensitivity and specificity values across these subgroups remained within a narrow range around the overall estimates.

**Figure 4.**
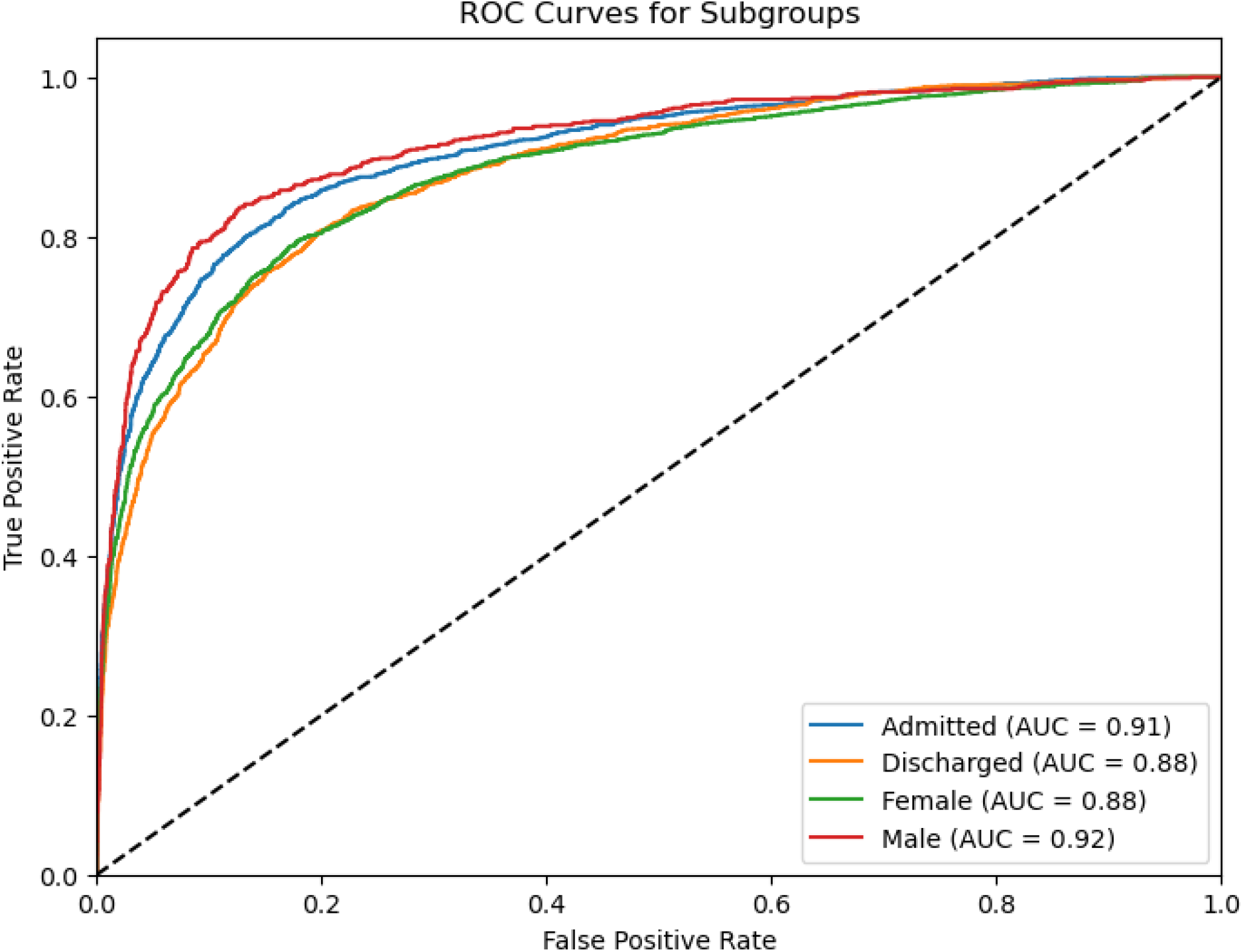
ROC curves for XGBoost model performance in patient subgroups (admitted, discharged, female, and male).

When evaluated against proxy measures of clinician decision-making, including documented UTI diagnoses and UTI-directed antibiotic prescribing, the XGBoost model yielded higher AUC values for culture-confirmed UTI than these benchmarks. Detailed numeric comparisons are provided in Supplementary Material.

## 4. Discussion

In this study, we developed and evaluated an interpretable machine learning framework for UTI prediction in emergency department patients using a large, multi-institutional dataset that has previously been used to develop predictive models [2,3]. Across four algorithms, XGBoost achieved the highest discrimination for general UTI prediction, with an AUC of 0.90, while Random Forest and Logistic Regression both achieved AUC values of 0.89 and the Decision Tree model an AUC of 0.86. The modest performance margin between XGBoost and logistic regression indicates that much of the predictive signal arises from relatively straightforward relationships, which may facilitate clinician understanding.

The reduced-feature XGBoost model performed similarly to the full 217-feature implementation, indicating that a small subset of 20 carefully selected variables captures the majority of predictive information. This parsimony is important for clinical implementation, where access to complete high-dimensional data may be limited and model simplicity supports robustness and transparency.

For complicated UTI prediction, the XGBoost model achieved an AUC of 0.97. This substantially higher performance compared to general UTI prediction suggests that complicated cases present with more distinctive clinical and laboratory patterns that can be captured by the model. High discrimination in this subgroup is clinically meaningful because complicated UTIs require more aggressive management, including broader-spectrum antibiotics and a lower threshold for admission.

Subgroup analyses showed that model performance was consistent across patient groups, with AUC values from 0.88 to 0.92. Performance was slightly higher among male patients and admitted patients, which may reflect differences in baseline risk and severity at presentation. Importantly, the model outperformed proxy clinical benchmarks based on discharge diagnoses and UTI-directed antibiotic prescribing, supporting the potential for data-driven decision support to improve on existing practice.

A central contribution of this work is the explicit focus on interpretability. The SHAP framework revealed that urinalysis components, particularly nitrite presence, bacterial counts, leukocyte esterase, and urinary white blood cells, dominate the model’s decision-making, which is consistent with established clinical reasoning for UTI diagnosis. This alignment between algorithmic and clinical reasoning is important for fostering appropriate trust[9].

Interpretability in this framework operates at both global and individual levels. At the global level, SHAP summary plots highlight the most influential features and their direction of effect, allowing clinicians to assess whether the model appears to rely on clinically plausible predictors. At the individual level, SHAP values for a single patient can be used to explain why a given predicted probability is high or low, by displaying how each feature contributed to the final output.

Our findings also highlight that not all influential features are direct biological markers. The presence of “not reported” laboratory variables among important predictors indicates that test-ordering patterns and documentation behaviours carry predictive information. This underscores the need for clinical expertise when interpreting machine learning outputs, as statistical associations do not necessarily imply causal mechanisms.

By coupling high discrimination with transparent explanations, the proposed framework supports a model of clinical integration in which predictions are used as an adjunct to, rather than a replacement for, clinician judgment. When model and clinician assessments are concordant, the tool may increase diagnostic confidence. When they diverge, SHAP-based explanations can prompt structured reflection about potential overlooked information, both in the data and at the bedside.

Our findings extend the foundational investigation by Taylor et al. (2018), who applied machine learning to the same ED dataset [5]. Their study demonstrated that an XGBoost model could achieve an AUC of approximately 0.904 for UTI prediction. We reproduced this level of discrimination using a comparable modelling strategy, with XGBoost achieving an AUC of 0.90 in our implementation. This agreement supports the robustness of performance estimates for this dataset.

Beyond reproducing overall performance, our work provides several methodological and conceptual extensions that differentiate it from the original analysis, despite the shared data source. First, we introduced a comprehensive explainability layer using SHAP, moving beyond reporting feature coefficients or importance scores to a unified framework for global and case-level explanations. This directly addresses the interpretability gap highlighted in previous work and responds to the demand for transparent AI in clinical settings[10]

Second, we systematically evaluated a reduced-feature XGBoost model that retained performance comparable to the full model. This parsimonious model is more amenable to implementation in settings where only a subset of variables is routinely available or where simplified models are preferred for operational reasons.

Third, we provided a detailed subgroup analysis by gender, disposition, and UTI complexity, as well as an explicit analysis of complicated UTIs. The exceptional performance for complicated UTI prediction and the relatively uniform performance across subgroups were not extensively characterized in previous studies.

More broadly, our work aligns with a growing literature on infection prediction and explainable AI in healthcare, and it illustrates how existing open datasets can be reanalyzed to develop models that are not only accurate, but also interpretable and implementation-focused [1–4].

### Study Limitations

Several limitations should be considered when interpreting these findings. First, the dataset derives from 2013 to 2016 encounters at four academic centres. Changes in antimicrobial resistance patterns, empiric prescribing practices, clinical testing protocols, and patient populations over time may limit contemporary applicability. External validation using more recent data from diverse practice settings is necessary before implementation.

Second, SHAP-based explanations describe how the model uses the available data, but they do not distinguish causal from non-causal associations. Important features may reflect confounding or institutional practice patterns that would not generalize. Clinical interpretation of feature attributions must therefore be cautious and contextual.

Finally, the outcome definition relies on urine culture as the gold standard[2]. Culture sensitivity is imperfect, particularly when antibiotics are given before sample collection, so some apparent model misclassifications may represent limitations of the reference standard rather than true errors.

### Future research directions

Several directions emerge from this work. Prospective clinical validation is essential. Real-time implementation studies should assess the impact of model use on antibiotic prescribing patterns, diagnostic accuracy, patient outcomes, and resource use. Randomized or quasi-experimental designs would provide the strongest evidence for clinical utility.

External validation across different healthcare systems, including community and rural hospitals and international sites, is needed to assess generalizability and to determine whether recalibration is necessary. Temporal updating using more recent data would allow the model to adapt to evolving UTI epidemiology and prescribing patterns.

Implementation science research should explore clinician perspectives on explainable AI for UTI diagnosis, preferred formats for explanations, and how to embed decision support tools into ED workflows without increasing cognitive burden. Comparative effectiveness studies could also evaluate the performance and usability of this framework relative to more traditional decision rules or newer rapid diagnostic technologies.

The high discrimination for complicated UTI suggests a potential role in risk stratification, identifying patients who may benefit from early broad-spectrum therapy, urological evaluation, or inpatient management. From an antimicrobial stewardship perspective, the model may help to reduce unnecessary antibiotic exposure in culture-negative patients by providing a structured, data-driven counterweight to overtreatment tendencies.

At a systems level, aggregate predictions and explanations could be used for quality improvement, helping to identify patterns of overdiagnosis and underdiagnosis and informing targeted educational interventions. The framework could also serve as an educational tool for trainees, by highlighting cases in which model predictions and clinical decisions diverge and prompting reflection on underlying reasoning.

## Conclusion

This investigation developed and validated an interpretable machine learning framework for urinary tract infection prediction in emergency department settings, using a large publicly available dataset previously analyzed with primarily black-box methods. XGBoost achieved robust discrimination for general UTI prediction and particularly strong performance for complicated UTI, and a reduced-feature model maintained comparable performance. SHAP-based explanations provided transparent feature attribution at both global and individual levels.

Translation from statistical performance to real-world impact will require prospective and external validation, careful implementation design, and engagement with clinicians and other stakeholders. Nonetheless, this study provides a foundation for integrating explainable machine learning into ED UTI diagnosis and illustrates how reanalysis of existing datasets can yield models that are better aligned with the needs of clinical practice.

## Data Availability

The link to the data has been provided as supplementary material in the manuscript

